# Global between-countries variance in SARS-CoV-2 mortality is driven by reported prevalence, age distribution, and case detection rate

**DOI:** 10.1101/2020.05.28.20114934

**Authors:** Haris Babačić, Janne Lehtiö, Maria Pernemalm

## Abstract

**Objective:** To explain the global between-countries variance in number of deaths per million citizens (*nD*_pm_) and case fatality rate (*CFR*) due to severe acute respiratory syndrome coronavirus 2 (SARS-CoV-2) infection.

**Design:** Systematic analysis.

**Data sources:** Worldometer, European Centre for Disease Prevention and Control, United Nations

**Main outcome measures:** The explanators of *nD_pm_* and *CFR* were mathematically hypothesised and tested on publicly-available data from 88 countries with linear regression models on May 1^st^ 2020. The derived explanators – age-adjusted infection fatality rate (*IFR_ad_j*) and case detection rate (*CDR*) – were estimated for each country based on a SARS-CoV-2 model of China. The accuracy and agreement of the models with observed data was assessed with *R*^2^ and Bland-Altman plots, respectively. Sensitivity analyses involved removal of outliers and testing the models at five retrospective and four prospective time points.

**Results:** Globally, *IFR_adj_* estimates varied between countries, ranging from below 0.2% in the youngest nations, to above 1.3% in Portugal, Greece, Italy, and Japan. The median estimated global *CDR* of SARS-CoV-2 infections on April 16^th^ 2020 was 12.9%, suggesting that most of the countries have a much higher number of cases than reported.

At least 93% and up to 99% of the variance in *nD_pm_* was explained by reported prevalence expressed as cases per million citizens (*nC*_pm_), *IFR_adj_*, and *CDR. IFR_ad_j* and *CDR* accounted for up to 97% of the variance in *CFR*, but this model was less reliable than the *nD_pm_* model, being sensitive to outliers (*R*^2^ as low as 67.5%).

**Conclusions:** The current differences in SARS-CoV-2 mortality between countries are driven mainly by reported prevalence of infections, age distribution, and *CDR*. The *nD_pm_* might be a more stable estimate than *CFR* in comparing mortality burden between countries.

## Introduction

The severe acute respiratory syndrome coronavirus 2 (SARS-CoV-2) has substantially affected the lives of billions of people.(1,2) An ongoing question in the public is how high is the direct mortality caused by SARS-CoV-2. Observations in the case fatality rate (*CFR*), i.e. the proportion of individuals with a confirmed SARS-CoV-2 infection who die, has raised concerns due to the high variability between countries, ranging from below 0.1% in Qatar and Singapore to above 15% in Belgium and France, on May 27^th^ 2020.(3,4)

The global average case detection rate was age (*CDR*) on March 30^th^ was estimated at 9%, suggesting that the true prevalence of infections is likely underestimated in most of the countries.(9) Studies suggest that the reported number of cases per million citizens (*nC_pm_*) is probably lower than the true number of infected individuals, and that this contributes to the varying *CFR* between countries.(5,6) *CFR* appears higher than the true infection fatality rate (*IFR*), i.e. the true proportion of individuals with a SARS-CoV-2 infection who will die in the population regardless of whether they are confirmed or not.(7) This was observed in China where the crude *CFR* estimate was 3.67%, whereas the age-adjusted overall *IFR* (*IFR_ad_j*) was estimated at 0.66%. (8)

The number of confirmed deaths per million citizens (*nD*_pm_) is a population-normalised measure of mortality used to compare countries. However, the varying *nD_pm_* in countries with similar *nC_pm_*, population size and similar mitigation strategies has also raised fears of potential varying virulence of the virus and different treatment capacity between countries. A recent multivariable model could explain only 62.5% of SARS-CoV-2 mortality variance between countries.(10) Explaining the remaining variance of the reported mortality as *nD_pm_* and *CFR* is extremely relevant for both the medical community and the public, to address public concerns. Furthermore, it is important to assess whether the adjusted mortality differs substantially between countries, in order to track the success of different strategies.

The aim of this study was to test two mathematical hypotheses that explain the global between-countries variance in SARS-CoV-2 mortality expressed as *nD_pm_* and *CFR* on real data.

## Methods

### Data sources

Global data on cumulative number of cases (*nC*), cumulative number of deaths (*nD*), cumulative number of tests (*nT*), number of tests per million citizens (*nT*_pm_), number of cases per million citizens (*nC*_pm_), and *nD_pm_* per country were downloaded from Worldometer.(3) Global data on number of new cases and deaths per day were downloaded from the European Centre for Disease Prevention and Control (ECDC).(4) Global data on age distribution and 2018 GDP per country were obtained from United Nations (UN) statistics.(11,12)

### Derivation of hypotheses

The overall *IFR_ad_j* per country estimated and weighed per nine groups, following the equation:

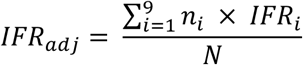

where *IFR_adj_* is the *IFR_adj_* in percentages (%), *N* is the total population size, *n_i_* is the number of susceptible individuals within an age group, *IFR_i_* is the *IFR* for that age group in % as estimated by Verity and colleagues.(8) For the purposes of this study, the *IFR_adj_* serves as an age-adjustment factor.

The *CDR* per country was estimated as the percentage of the estimated cases that have been confirmed cases, following the approach of Vollmer & Bommer (9):

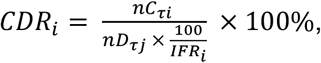

where *CDRi* is *CDR* in %, *IFR_i_* is *IFR_ad_j* in %, *nC_τi_* is cumulative number of confirmed cases at time *τi*, and *nD_τj_* is cumulative number of confirmed deaths at time *τj*. Following the Verity model(8), *τi* is 14 days before *τj* in this approach, based on the estimate that on average 18.8 days pass from the onset of symptoms to death, holding a conservative assumption that on average 4.8 days pass from symptom onset to case detection.

From these equations, *nD_τj_* is implied to have an inverse relation with the *CDRi* and will depend on the cumulative number of cases 14 days before the *nD_τj_* have occurred, and the age-adjusted *IFR_i_*:

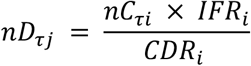

Assuming that the number of cases at the time of *nD_τj_* (*nC_τj_*) will have a constant dependence on the *nC_τi_*, as observed repeatedly in epidemics, including SARS-CoV-2, *nC_τj_* can replace it in the equation. In order to explain the population-normalised number of deaths – *nD_pm_*, one has to normalise the *nC_τj_* per population size with *nC_pm_*, deriving that:

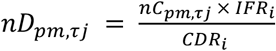

where *nD_pm_* at time *τj* will be higher in countries with higher *nC_pm_* at time *τj* and higher *IFR_adj_*, whereas it will be lower in countries with the same *nC_pm_* and *IFR_adj_* that have higher *CDR*.

Following that the *CFR* is the proportion of *nD_τj_* from the *nC_τj_*, the subsequent relationship between the *CFR*, the *IFR_adj_* and *CDR* is implied as:

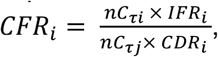

where *CFR_t_* is the *CFR* of a country, *IFR_t_* is the *IFR_adj_* of a country, *nC_τj_* is the cumulative number of cases on the day of *nD_τj_>*, and *nC_τi_* is the cumulative number of cases two weeks before *τj*. Again, assuming that *nC_τj_* will have a constant dependence on the *nC_τi_*, they can be omitted from the equation, deriving that:

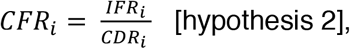

where the *CFR* will be higher in countries with higher *IFR_adj_* and will have an inverse relation with the *CDR*. Hypothesis 2 implies that older countries will have higher *CFR* and countries with higher *CDR* will have lower *CFR*, and predicts that the *nC_τj_* will not drive *CFR*.

### Testing hypotheses on real data

To test hypothesis 1, we built linear regression model 1 (*nD_pm_* model), to explain *nD_pm_* with *nC_pm_, IFR_ad_j*, and *CDR*. To test hypothesis 2, we built linear regression model 2 (*CFR* model), to explain *CFR* with *IFR_adj_* and *CDR*. Only countries with more than 1,000 cases were included in the analyses. All variables were normalised by log transformation. We have additionally tested whether GDP, *nT_pm_*, and duration of epidemic (as days from first case) could explain the mortality after being added to the models.(10) The accuracy was assessed with *R^2^*, and the agreement was analysed graphically with the Bland-Altman mean difference plot.(13)

*Sensitivity analyses*. To address uncertainty, we removed outliers outside of the 95% confidence intervals (95% CI) of the Bland-Altman plots, and reiterated the analyses retrospectively on April 4^th^, 8^th^, 12^th^, 21^st^, 24^th^, and prospectively on May 7^th^, 11^th^, 18^th^, and 27^th^ 2020.

### Data and code availability

The study is conducted according to the Guidelines for Accurate and Transparent Health Estimates Reporting.(14) The code, data, and results are publicly available at https://aithub.com/harbab/covid_19mortality. All analyses were performed in R V.3.6.1.

## Results

### Descriptive statistics

As of May 1^st^ 2020, a total of 214 countries in the world have reported SARS-CoV-2 infections. Of these, 88 countries have reported more than 1,000 SARS-CoV-2 infections. The estimated *IFR_adj_* varied from below 0.2% in the youngest nations of Ivory Coast, Guinea, Nigeria, UAE, Cameroon, and Afghanistan, up to above 1.3% in the world’s oldest nations of Germany, Portugal, Greece, Italy, and highest in Japan with 1.6%. The global average *CDR* on April 16^th^ 2020 was 22.12% (median: 12.9%, SD: 32.47), suggesting that most of the cases were undetected. Only two countries detected more than 100% of expected cases – Iceland (154.50%) and Singapore (234.95%). Estimates for each country are shown in Table S1, supplementary information.

### Model 1 – *nDp_m_*

Univariate analyses showed that *nC_pm_, IFR_adj_*, and *nT_p_*_*m*_ could explain 65.57%, 40.29%, and 25.91% of the variance in *nD_pm_*, respectively (p < 3.922^−07^). The *CDR* was not univariately associated with *nD_pm_* (p = 0.738). However, combined together *nC_P_m*, *IFR_adj_* and *CDR* could explain 97.18% of the variance in *nD_p_*_*m*_ (p < 2.2^−16^). Introducing *nT_p_*_*m*_ to the model only slightly improved the *R*^*2*^ to 0.9728 (p < 2.2^−16^). All four variables were included in the final model 1 (Table 1). The relationship between the variables was as hypothesised mathematically. The model showed almost perfect accuracy (Figure 1A) and high agreement (Figure 1B) in explaining *nD_pm_*. Some countries were outliers (Figure 1C).

**Table 1.** Linear model 1 – explaining the log-transformed citizens *nD_pm_*. The model had a residual standard error of 0.3073 on 79 degrees of freedom (4 observations with missing data). Multiple *R*^2^ = 0.9741, adjusted *R*^2^ = 0.9728. F-statistic: 743.8 on 4 and 79 DF, p-value: < 2.2^−16^.

| Coefficient | Estimate | 95% CI | Std. error | t-value | p-value | significance |
| --- | --- | --- | --- | --- | --- | --- |
| Intercept | -0.9277 | -1.3786, -0.4769 | 0.227 | -4.096 | 0.0001 | *** |
| log ($IFR_{adj}$) | 1.2951 | 1.1656, 1.4246 | 0.065 | 19.907 | 1.61<sup>-32</sup> | *** |
| log ($nC_{pm}$) | 0.9432 | 0.8647, 1.0216 | 0.039 | 23.931 | 6.03<sup>-38</sup> | *** |
| log ($CDR$) | -0.9899 | -1.0785, -0.9012 | 0.045 | -22.218 | 1.01<sup>-35</sup> | *** |
| log ($nT_{pm}$) | 0.0922 | 0.0071, 0.1774 | 0.043 | 2.156 | 0.0341 | * |

\*\*\*, statistically highly significant; \*, statistically low significance.

**Figure 1.**
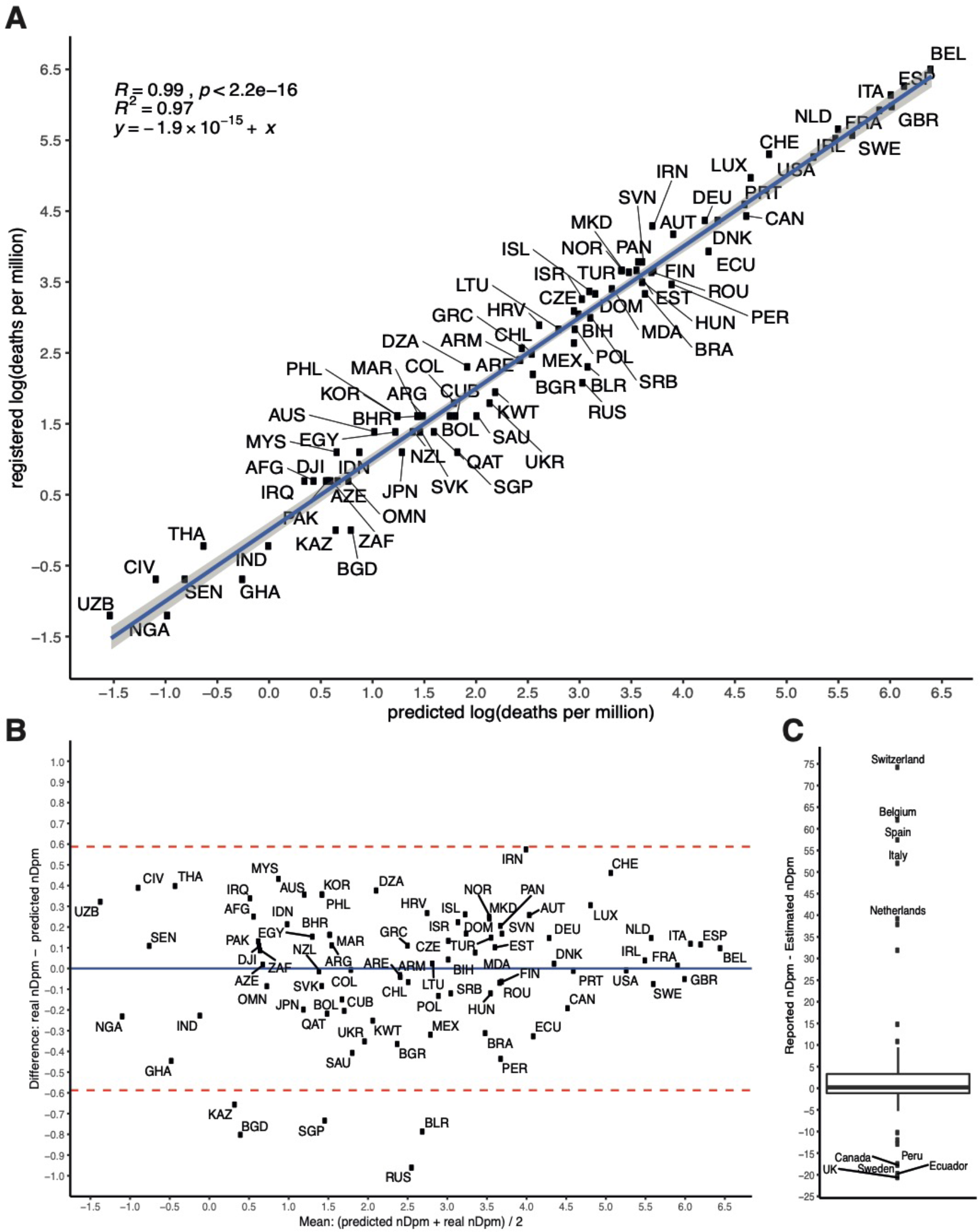
Agreement between predicted and observed deaths per million citizens (*nD_pm_*). The countries are annotated with their country code. **A.** Predicted log-normalised *nD_pm_*(x axis) vs log-normalised observed *nD_pm_* (y axis). The model could almost perfectly predict the *nD_pm_* in a linear fashion. The blue line is model fit and the shades are 95% Cl; **B**. Bland-Altman plot. The mean of the predicted log-normalised *nD_pm_* and observed log-normalised *nD_pm_* is plotted on the x axis, whereas the difference on a log scale between the observed *nD_pm_* and predicted *nD_pm_* is plotted on the y axis. The mean difference between the observed *nD_pm_* and predicted *nD_pm_* was 0 (blue, full line), with the 95% confidence intervals (red, dashed lines) containing most of the values. Five countries were outliers in this model, having less *nD_pm_* than predicted: Russia, Belarus, Singapore, Bangladesh, and Kazakhstan; C. Countries outliers. Actual difference between observed *nD_pm_* and predicted *nD_pm_* in numbers. The labelled countries in the upper part of boxplot (>95^th^ quantile) had much more observed *nD_pm_* than predicted, whereas the labelled countries in the lower part had much less *nD_pm_* than predicted by the model.

Univariately, *nD_pm_* had a positive association with GDP (p = 8.213^−11^) and the duration of the epidemic (0.0227). Plowever, neither variable had an association with nD_pm_when added to model 1 (p = 0.308 and 0.196, respectively). GDP had a positive correlation with all four explanators of model 1 (p < 9.341^−06^), most evidently with *nT_pm_*(*R*^2^ = 0.592), and was thus redundant in the model. GDP and duration of epidemic are possibly unstable explanators that can be useful in stratified analyses per continents and regions.

### Model 2 – *CFR*

Univariately, *IFR_ad_j* and *CDR* explained 17.68% and 40.35% of the variance in *CFR*, respectively. When combined together, *IFR_ad_j* and *CDR* accounted for 91.84% of the variance in *CFR* (Table 2). The predicted *CFR* also had high accuracy and high agreement with observed *CFR* (Figure 2). Both *nC_pm_* and *nC_τj_* were not associated with the *CFR* when added to model 2 independently, confirming the assumption on which hypothesis 2 relies. None of the additional variables (*nT*_pm_, GDP, or duration of epidemic) was associated with *CFR* univariately or when added to model 2. *nT_pm_* and GDP were also not associated with *CFR* in a previous report (10).

**Table 2.**
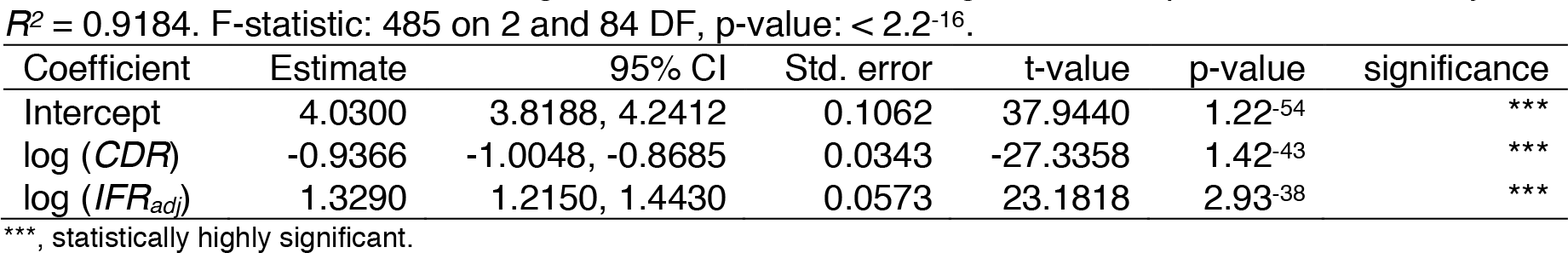
Linear model 2 – explaining the log-transformed *CFR*. The model had a residual standard error of 0.3126 on 84 degrees of freedom (1 missing data). Multiple *R*^2^ = 0.9203, adjusted *R*^2^ = 0.9184. F-statistic: 485 on 2 and 84 DF, p-value: < 2.2^−16^.

| Coefficient | Estimate | 95% CI | Std. error | t-value | p-value | significance |
| --- | --- | --- | --- | --- | --- | --- |
| Intercept | 4.0300 | 3.8188, 4.2412 | 0.1062 | 37.9440 | 1.22<sup>-54</sup> | *** |
| log ($CDR$) | -0.9366 | -1.0048, -0.8685 | 0.0343 | -27.3358 | 1.42<sup>-43</sup> | *** |
| log ($IFR_{adj}$) | 1.3290 | 1.2150, 1.4430 | 0.0573 | 23.1818 | 2.93<sup>-38</sup> | *** |

\*\*\*, statistically highly significant.

**Figure 2.**
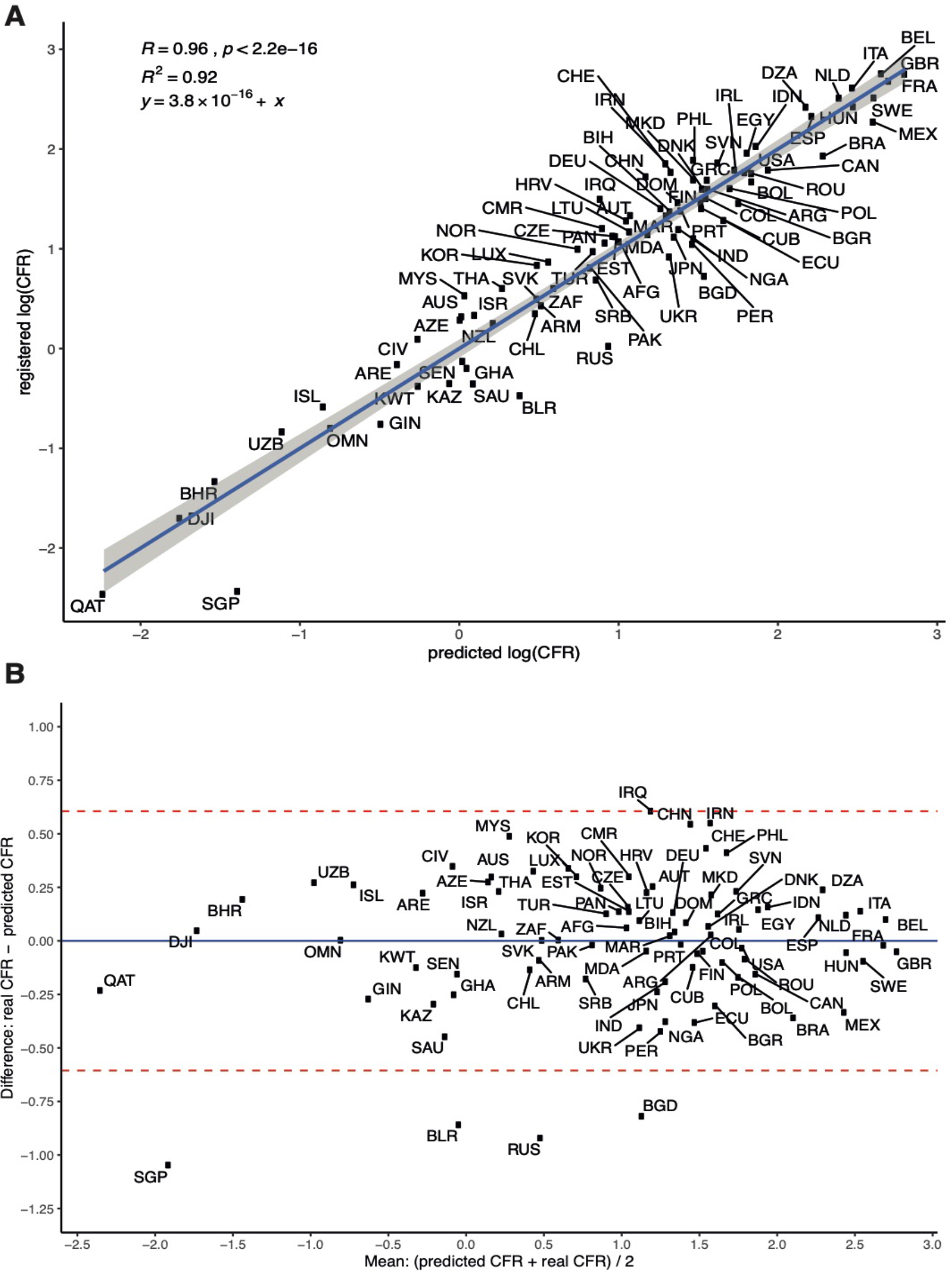
Agreement between observed and predicted case fatality rate *(CFR)*. The countries are annotated with their country code. **A.** Predicted log-normalised *CFR*(x axis) vs log-normalised observed *CFR*(y axis). The model could predict almost perfectly the *CFR* in a linear fashion. The blue line is model fit and the shades are 95% CI; **B**. Bland-Altman plot. The mean of the predicted log-normalised *CFR* and observed log-normalised *CFR* is plotted on the x axis, whereas the difference on a log scale between the observed *CFR* and predicted *CFR* is plotted on the y axis. The mean difference between the observed *CFR* and predicted *CFR* was 0 (blue, full line), with the 95% confidence intervals (red, dashed lines) containing most of the values. Four countries were outliers in this model, having lower *CFR* than predicted: Russia, Belarus, Singapore, and Bangladesh.

### Sensitivity analyses

Reiterating the analysis at five retrospective and four prospective time-points showed that model 1 could robustly explain at least 93% of the variance in *nD_pm_* (at least 95% after removing outliers), but model 2 had lower accuracy at earlier stages of the pandemic (Figure 3). Less countries had > 1,000 cases at earlier time-points (range: 56 on April 4^th^ – 110 on May 27^th^). The *nT_pm_* was an unreliable explanator of *nD_pm_* that accounted for a very small proportion of variance that can be omitted; the effect of *nTpm* is possibly underestimated due to its association with *nC_pm_*.

**Figure 3.**
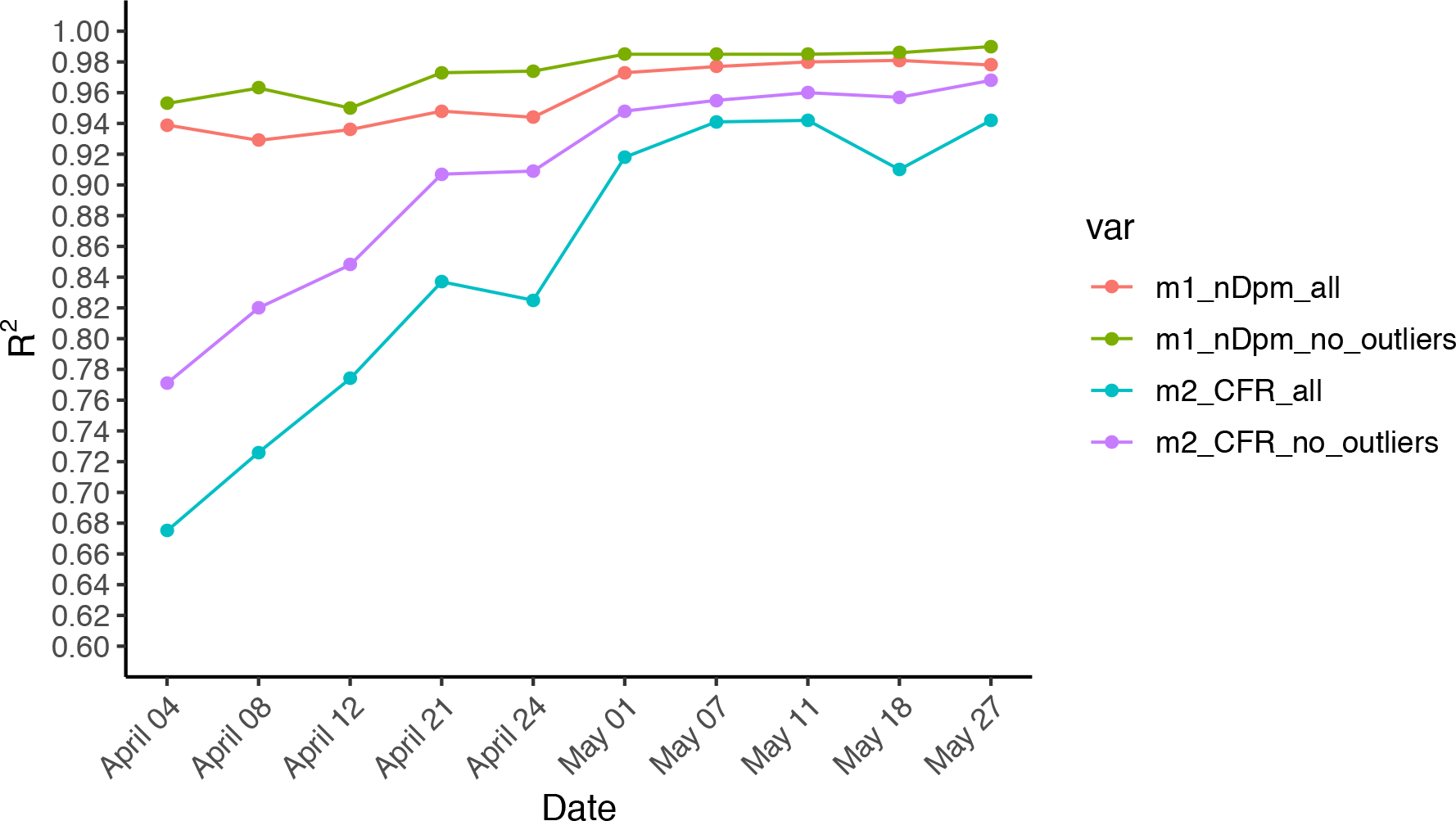
*R^2^* at ten time points of linear model 1 (explaining *nD_pm_)* and linear model 2 (explaining *CFR*). m1_nDpm_all, model 1 with all countries included; m1_nDpm_no outliers, model 1 after excluding outliers; m2_CFR_all, model 2 with all countries included; m2_CFR_no_outliers, model 2 after excluding outliers.

The *CFR* model was more sensitive to outliers compared to the *nD_pm_* model, with a higher average decrease in *R*^2^ of 5.53% (range: 1.4–9.6%, median: 5.85%) compared to an average decrease of 1.59% for the *nD_pm_* model (range: 0.5–3.4%, median: 1.2%) when including outliers (p = 0.0035). The assumption of no effect of *nC_τj_* on *CFR* was violated at some time-points.

On average, several countries had > 25 more *nD_pm_* than expected – Italy, Belgium, Switzerland, Spain, the Netherlands, and Iran, whereas others had on average > 5 less *nDpm* than expected – UK, Peru, Brazil, Belarus, Russia, Canada, Chile and Kuwait. Likewise, Italy, Algeria, Iran, the Netherlands, China, Belgium, Iraq, Indonesia, Spain, Switzerland, and the Philippines had on average > 1.5% higher *CFR* than expected according to the model, whereas Bangladesh, Ukraine, Brazil, Bolivia, Mexico, Belarus, Russia, Peru, and Honduras had on average > 1 % lower *CFR* than expected.

At prospective time-points in May, three countries had consistently higher than 100% *CDR*, reporting more cases than expected: Singapore (range: 404–854%), Iceland (range: 159–160%), and Qatar (range: 121–210%). Detailed results from the sensitivity analyses are available in supplementary information.

## Discussion

Most of the global variance in *nD_pm_* between countries was explained by reported prevalence of SARS-CoV-2 infections (*nC*_pm_) and age distribution as expressed with the *IFR_adj_*. This has to be further adjusted for the *CDR*, which has an inverse relation with the *nD_pm_*, but only in the context of using *nC_pm_* and *IFR_ad_j* to explain *nD_pm_*. As expected, the richer countries were better at testing and detecting cases, but were also older and had a higher infection mortality burden. The *CFR* is also dependent on the *IFR_adj_* and the *CDR*, but does not depend on the prevalence or the total number of SARS-CoV-2 confirmed cases.

Some countries remain outliers, having consistently higher or lower mortality than expected according to the models. This is possibly due to consistent misreporting (10), differences in reporting deaths, diagnostic bias, sex distribution and average age of individuals who died – countries with on average higher mortality than expected possibly had more older people and more men infected and dying.(15)

The observation that several countries have detected a higher number of cases than expected and had lower observed *CFR* than *IFR_adj_* (see supplementary information), supports the notion that the current *IFR* is overestimated and that the actual mortality is lower than estimated. To confirm these observations, serological surveys of populations will be essential in correctly estimating the true prevalence and mortality of SARS-CoV-2.

### Strengths and limitations

These models use very few explanators while maintaining high accuracy in explaining mortality. The sensitivity analyses demonstrated the robustness of the mathematical models when tested on real data. There is a remaining small proportion of variance than cannot be explained by the models, and this can be due to data mishandling or estimation errors, which limit the study. Independent of these limitations, the *nD_pm_* model remained robust. The *CFR* model was more sensitive to outliers than the *nD_pm_* model, and might be a less stable mortality outcome to follow SARS-CoV-2 mortality burden over time and across countries. The models were somewhat less accurate at earlier stages, which can be due to the amount of data (number of countries) used to build the models.

### Conclusions and policy implications

Overall, this study demonstrates that most countries are on a similar SARS-CoV-2 mortality trajectory as the number of cases increases, after adjusting for age distribution and *CDR*. These models should be used for less biased comparisons of mortality between countries. The *nD_pm_* model appears as a more stable indicator of SARS-CoV-2 infection mortality burden and should be favoured in following and comparing mortality within and between countries.

### Evidence before this study

-Verity and colleagues (Lancet Inf Dis 2020) have estimated the SARS-CoV-2 infection fatality rates *(IFR)* per age groups, and Vollmer & Bommer (2020) have estimated that the average case detection rate *(CDR)* of SARS-CoV-2 infections in 40 countries was below 10% end of March.

-No studies have been published explaining the global SARS-CoV-2 variance in mortality. A medRxiv preprint by Shagam (2020) reports that approximately 60% of SARS-CoV-2 mortality variance can be explained by gross domestic product per capita in United States dollars (GDP), latitude, hemisphere, press freedom, population density, fraction of citizens over 65 years old, and outbreak duration.

### Added value of this study

-The models in this study demonstrate that most of the between-countries variance in SARS-CoV-2 mortality can be explained with two to three explanators, maintaining high accuracy. This can help to alleviate public concerns of potential varying virulence of the virus, and provide a less biased, standardised comparison of mortality burden between countries.

-In the setting of lacking an effective and safe treatment and/or vaccine against SARS-CoV-2, most of the countries will be on a similar SARS-CoV-2 mortality trajectory as the number of cases increases, after adjusting for the age distribution of the population and the case detection rate.

## Data Availability

The code, data, and results will be publicly available upon publication. All analyses were performed in R V.3.6.1.

https://github.com/harbab/covid_19_mortality

## Acknowledgments

We express gratitude to Dr. Petter Brodin, Dr. loannis Siavelis, and Dr. Emilie Friberg for reading the draft and providing fruitful feedback.

## Patient and public involvement

The study is conducted with publicly available data, and does not include individual patient or public involvement.

## Ethical standards

The study was performed according to the ethical standards expressed in the Declaration of Helsinki. This study does not require ethical approval.

## Contribution

HB designed the study, derived the hypotheses, collected, analysed and interpreted the data, and wrote and edited the manuscript. JL and MP assisted in design and interpretation of the study, supervised the work, reviewed and edited the manuscript.

## Transparency

The corresponding author (HB) affirms that this manuscript is an honest, accurate, and transparent account of the study being reported; that no important aspects of the study have been omitted; and that any discrepancies from the study as planned have been explained.

## Dissemination

not applicable.

## References

1. Studdert DM, Hall MA. Disease Control, Civil Liberties, and Mass Testing – Calibrating Restrictions during the Covid-19 Pandemic. N Engl J Med. 2020 Apr;

2. McKee M, Stuckler D. If the world fails to protect the economy, COVID-19 will damage health not just now but also in the future. Nat Med. 2020 Apr;

3. Worldometer. COVID-19 Coronavirus Pandemic [Internet]. 2020 [cited 2020 May 27]. Available from: https://www.worldometers.info/coronavirus/

4. European Centre for Disease Prevention and Control ECDC. Geographic distribution of COVID-19 cases worldwide [Internet]. [cited 2020 May 27]. Available from: https://www.ecdc.europa.eu/en/publications-data/download-todays-data-geographic-distribution-covid-19-cases-worldwide

5. Bendavid E, Mulaney B, Sood N, Shah S, Ling E, Bromley-Dulfano R, et al. COVID-19 Antibody Seroprevalence in Santa Clara County, California. medRxiv [Internet]. 2020 Jan 1;2020.04.14.20062463. Available from: http://medrxiv.org/content/early/2020/04/30/2020.04.14.20062463.abstract

6. Li R, Pei S, Chen B, Song Y, Zhang T, Yang W, et al. Substantial undocumented infection facilitates the rapid dissemination of novel coronavirus (SARS-CoV-2). Science. 2020 May;368(6490):489–93.

7. Baud D, Qi X, Nielsen-Saines K, Musso D, Pomar L, Favre G. Real estimates of mortality following COVID-19 infection. The Lancet. Infectious diseases. United States; 2020.

8. Verity R, Okell LC, Dorigatti I, Winskill P, Whittaker C, Imai N, et al. Estimates of the severity of coronavirus disease 2019: a model-based analysis. Lancet Infect Dis. 2020 Mar;

9. Vollmer S, Bommer C. Average detection rate of SARS-CoV-2 infections is estimated around nine percent [Internet]. 2020 [cited 2020 May 12]. Available from: https://www.uni-goettingen.de/en/606540.html

10. Shagam L. Untangling factors associated with country-specific COVID-19 incidence, mortality and case fatality rates during the first quarter of 2020. medRxiv [Internet]. 2020 Jan 1;2020.04.22.20075580. Available from: http://medrxiv.org/content/early/2020/04/27/2020.04.22.20075580.abstract

11. United Nations, Department of Economic and Social Affairs, Population Dynamics UN. World Population Prospects 2019, Online Edition. [Internet]. 2020 [cited 2020 Apr 12]. Available from: https://population.un.org/wpp/Download/Standard/Population/

12. UN Data, United Nations Statistics Division UN. Per capita GDP at current prices – US dollars [Internet]. 2020 [cited 2020 May 12]. Available from: https://data.un.org/Search.aspx?q=gdp

13. Bland JM, Altman DG. Statistical methods for assessing agreement between two methods of clinical measurement. Lancet (London, England). 1986 Feb;1(8476):307–10.

14. Stevens GA, Alkema L, Black RE, Boerma JT, Collins GS, Ezzati M, et al. Guidelines for Accurate and Transparent Health Estimates Reporting: the GATHER statement. Lancet (London, England). 2016 Dec;388(10062):e19–23.

15. Onder G, Rezza G, Brusaferro S. Case-Fatality Rate and Characteristics of Patients Dying in Relation to COVID-19 in Italy. JAMA. 2020 Mar;

